# Development of a COVID-19 risk-assessment model for participants at an outdoor music festival: Evaluation of the validity and control measure effectiveness

**DOI:** 10.1101/2022.02.28.22271676

**Authors:** Michio Murakami, Tsukasa Fujita, Pinqi Li, Seiya Imoto, Tetsuo Yasutaka

## Abstract

We developed an environmental exposure model to estimate the Coronavirus Disease 2019 (COVID-19) risk among participants at an outdoor music festival and validated the model using a real cluster outbreak case. Furthermore, we evaluated the extent to which the risk could be reduced by additional infection control measures such as negative proofs of antigen tests on the day of the event, wearing masks, disinfection of environmental surfaces, and vaccination. The total number of already- and newly-infected individuals who participated in the event according to the new model was 47.0 (95% uncertainty interval: 12.5–185.5), which is in good agreement with the reported value (45). Among the additional control measures, vaccination, mask-wearing, and disinfection of surfaces were determined to be effective. Based on the combination of all measures, a 94% risk reduction could be achieved. In addition to setting a benchmark for an acceptable number of newly-infected individuals at the time of an event, the application of this model will enable us to determine whether it is necessary to implement additional measures, limit the number of participants, or refrain from holding an event.

## INTRODUCTION

During the global Coronavirus Disease 2019 (COVID-19) outbreak, the assessment and management of the infection risk during mass gatherings have become urgent issues ^1^. One risk assessment method is the epidemiological approach. To date, the COVID-19 infection risk related to events has been assessed using randomized controlled trials ^2^ or observational studies including both events with and without the use of measures such as mask-wearing ^3^. However, in the absence of infection control measures, participating in an event may result in a large number of infected individuals (i.e., clusters) ^4^. From an ethical perspective, having studies that actively use events without adequate control measures may not be suitable ^5^.

To overcome such limitations of existing studies, environmental exposure models may be applied and their effectiveness should be assessed. We previously developed an environmental exposure model to assess the COVID-19 risk among spectators at the opening ceremony of the Tokyo 2020 Olympic Games and evaluated the effectiveness of the implementation of control measures, including mask-wearing, physical distance, ventilation, disinfection, and handwashing ^6^. Additionally, we conducted parametric studies to evaluate the effects of the number of spectators, capacity proportions, and infection prevalence by extending the model to other sporting events ^7^. In another study, we evaluated the effects of vaccine-testing packages ^8^. We confirmed the validity of the model based on the fact that no newly-infected individuals were observed among the participants of professional baseball and soccer events in the fiscal year 2020 ^7^; however, this validation has limitations due to the unavailability of active testing after these events.

Therefore, in this study, we focused on a cluster outbreak case that occurred during an outdoor music festival event with inadequate infection control measures that was held in Japan during the emergence of the Delta variant. The objectives of this study were as follows: First, we extended the environmental exposure model to assess the COVID-19 risk at a music festival and validate the model by comparing the model estimates with the actual number of reported infected individuals. Second, we evaluated the reduction in the infection risk by applying the developed model to a hypothetical situation in which the event was held but additional or enhanced measures were implemented. This is the first study in which an environmental exposure model for the estimation of the infection risk was validated using a case study of an actual cluster outbreak among participants at a mass gathering event.

## METHODS

### Event and participants

The target event in this study was Namimonogatari2021, an outdoor Hip Hop festival held at the Aichi Sky Expo (3.5 ha) in the Aichi Prefecture from 9:00 to 21:00 (JST) on August 29, 2021 ^9^. The festival was held during the emergence of the Delta strain. In total, 7,392 people attended the festival and 45 infected individuals were reported ^9^. Of the participants, 1,154 were tested using the free polymerase chain reaction (PCR) tests that were conducted in the Aichi Prefecture and Nagoya City. As of September 13, a total of 658 test results were known and included eight positive cases. The reported number of infected cases (i.e., 45) included infected individuals identified in other areas ^10^. The number of infected people in the Aichi Prefecture during the week before this event (August 22– 28) was 12,072 ^11^. By dividing by the total population of the Aichi Prefecture ^12^, the number of infected persons per 10 million people was converted to 2,290 persons per day. Assuming that the proportion of asymptomatic individuals was 0.46 ^13^, the number of days between infectivity onset and recovery for asymptomatic individuals was 9.3 days and the number of days from infectivity onset to symptom onset for symptomatic individuals was 2.3 days ^14^. The crude probability of a participant being an infector (*P*_0_) is 1.3 × 10^−3^ based on weighting the infectivity time and the proportion of asymptomatic and symptomatic individuals ^6^. This number may be underestimated because several asymptomatic infectors may have not been identified.

### Model development

In this study, we extended a previously established model ^6-8^ to music festivals. In this model, the virus emission by asymptomatic infectors through talking, coughing, and sneezing is divided into four pathways: direct droplet spray, direct inhalation of inspirable particles, hand contact, and inhalation of respirable particles via air. The viral concentration was calculated after considering the inactivation in the environment and the risk of infection was estimated from several environmental and human behavioral parameters, including the breath volume and the frequency of hand contact with the surface, as well as the dose-response equation. The dose-response equation for the infection risk used in this study was based on the severe acute respiratory syndrome coronavirus (SARS-CoV) in mice ^15^ and the proportion of asymptomatic infected individuals in humans ^13^ because the equation was established on the basis of a wide range of doses. This parameter was similar to that for SARS-CoV-2 obtained from ferrets and the estimated human exposure ^16^. The estimated infection risk was slightly lower than the infection risk observed in the SARS-CoV-2 human challenge ^17^; the risk of infection at 55 focus forming unit was 53% in the human challenge, whereas it was 25% (95% uncertainty interval [UI]: 15–48) in this study.

The activities of the music festival participants were categorized into five behavioral patterns, that is, (A) attending live performances (60 min × 6 times); (B) entering, exiting, and resting (50 min × 6 times); (C) using restrooms (2 min × 3 times); (D) ordering at concession stands (1 min × 4 times); and (E) eating (25 min × 2 times); representing a total of 720 min. For each behavioral pattern, the amount of exposure was calculated according to the type of person exposed: (1) people accompanying the infector, (2) people in front of the infector at live performance venues, (3) peoples exposed in restrooms, (4) people exposed at concession stands, and (5) others. The types and numbers of people exposed are shown in Table S1 and the exposure pathways and doses for each behavioral pattern are shown in Tables S2 and S3.

By considering the actual size of the venue, number of spectators, and *P*_0_, we calculated the exposure related to the above-mentioned behavioral patterns. Considering the possibility that the Delta-variant strain has a 1,000-fold higher viral load than the wild-type strain ^18^, we carried out a sensitivity analysis and analyzed the results under conditions in which the concentration of the virus in saliva varied 10-, 100-, and 1,000-fold relative to the wild-type strain.

In the base scenario (without additional measures), mask-wearing and vaccination were considered. The amount of virus emitted by the infector differs depending on whether the infector wears a mask or not ^8^. Furthermore, exposed individuals wearing masks have a reduced frequency of contact with facial mucosal membranes ^6^. The mask-wearing proportions of the participants were set as follows: While the mask-wearing proportion among the Japanese public is extremely high (>85%) ^19^, the target event has been criticized for not ensuring that masks were worn ^9^. Therefore, we conducted a sensitivity analysis in which we assumed that 50% of the participants wore masks (base scenario) and then varied the mask-wearing proportion from 0% to 100% in 10% increments under conditions in which the Delta-variant concentration in saliva was 100-fold relative to the wild-type strain. The participants were divided into mask-wearers and non-wearers according to the mask-wearing proportion and the exposure dose was calculated for each category.

The percentage of people who received two doses of the vaccine was set to 45% based on the Japanese average ^20^. Considering that for many vaccinated individuals the elapsed time since the second vaccination was less than three months at the time of the event (two-dose vaccination coverage on May 29, 2021: 3% based on the Japanese average ^20^), the vaccine was assumed to be 80% effective in preventing infection with the Delta strain, which was emerging at the time of the event ^21^. The risk of infection in consideration of vaccination was assessed following a previous study ^8^.

Furthermore, in comparison to Supersonic (September 18–19, 2021) ^22^, which is known as an outdoor music festival with thorough infection control measures, we evaluated the risk of infection based on the addition of further hypothetical control measures under conditions in which the Delta-variant concentration was 100-fold relative to the wild-type strain:

a. Antigen testing: By conducting qualitative antigen testing for all participants on the day of the event, we reduced *P*_0_ by assuming that asymptomatic infectors who tested positive would be excluded from the event ^8^.
b. Distance: The distance from people during the entry, exit, and rest was set to 1.5 m and the distance from people during the attendance of live performances was set to 1 m. The number of people in front of the infector during the attendance of one live performance changed from three to one.
c. Mask-wearing: The mask-wearing proportion of the participants was set to 100%.
d. Restriction of talking during the attendance of live performances and meals: The frequency of talking during the attendance of live performances and meals was set to 0.03.
e. Disinfection: Disinfection after ordering at concession stands reduces the viral concentration on the surfaces to 1/1,000 ^6^.
f. Handwashing: Washing hands after using the restroom reduces the viral concentration on fingers to 1/100 ^6^.
g. Vaccination: The vaccination coverage of the participants was set to 100%. In this case, *P*_0_ did not change. This risk reduction may be underestimated because vaccinated individuals are considered to have a lower probability of being infected than unvaccinated individuals.
h. All measures (a–f) are implemented.
i. All measures (a–g) are implemented.

In addition, with measure (h) in place, analyses were conducted under conditions in which the number of participants or *P*_0_ was reduced from the base scenario to 75%, 50%, 25%, and 10%.

The model was run 10,000 times for each scenario.

## RESULTS

The total number of already- and newly-infected individuals, who participated in the event, was 24.8 (95% UI: 9.2–48.1), 47.0 (95% UI: 12.5–185.5), and 172.7 (95% UI: 25.1–610.0) for those with a 10-, 100-, and 1,000-fold increase in the Delta-variant viral concentrations relative to the wild-type strain, respectively (Figure 1). These results concur with the reported number of infected cases (45). Under a 100-fold viral concentration and mask-wearing proportion ranging from 0% to 100%, the total number of infected individuals varied from 73.0 (95%UI: 14.7–348.1) to 25.5 (95%UI: 9.6– 48.9; Figure S1).

**Figure 1.**
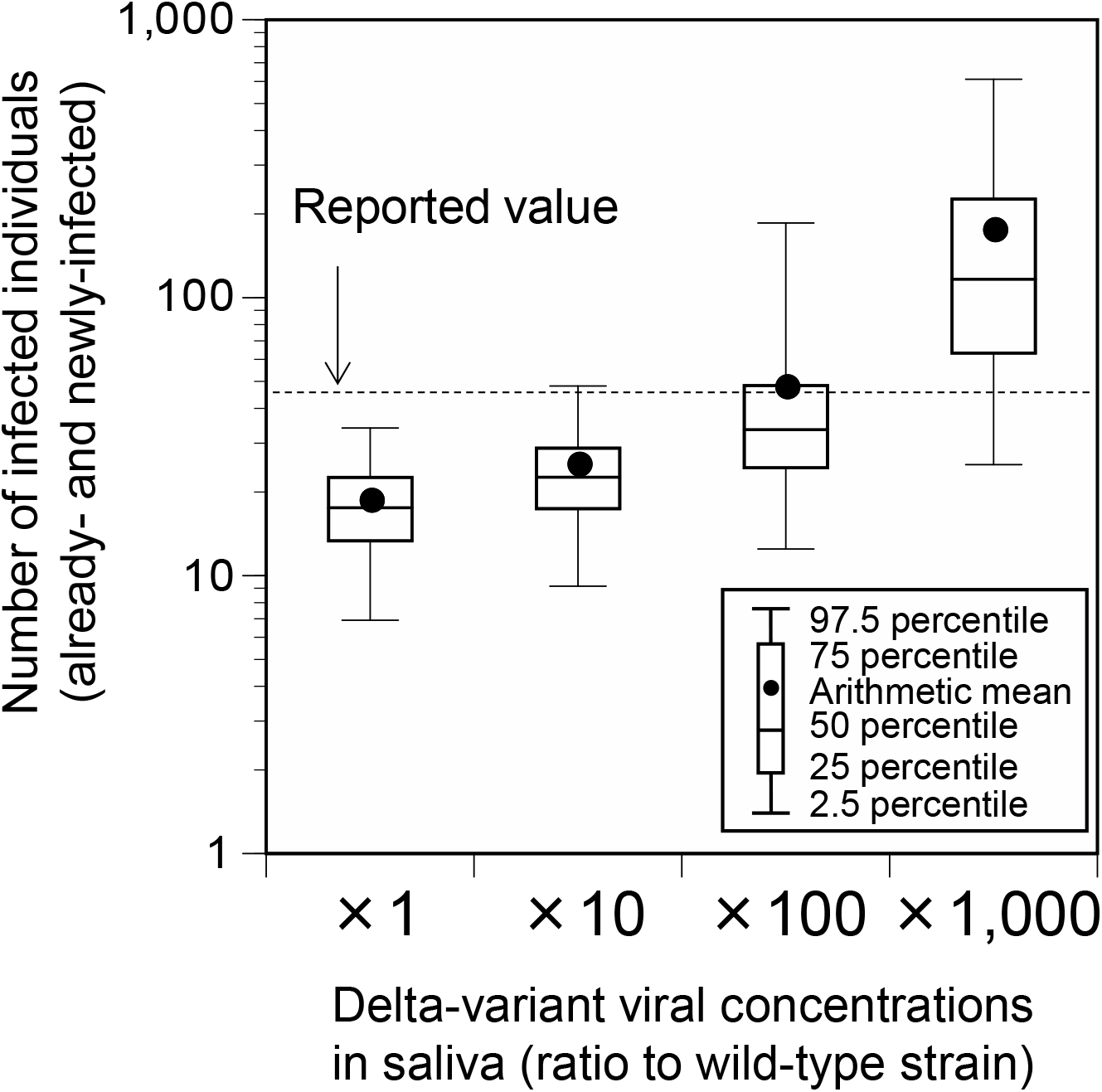
Comparison of the estimated and reported numbers of already- and newly-infected individuals (base scenario). Already-infected individuals represent those who were infectors at the time they participated in the event.

**Figure 2.**
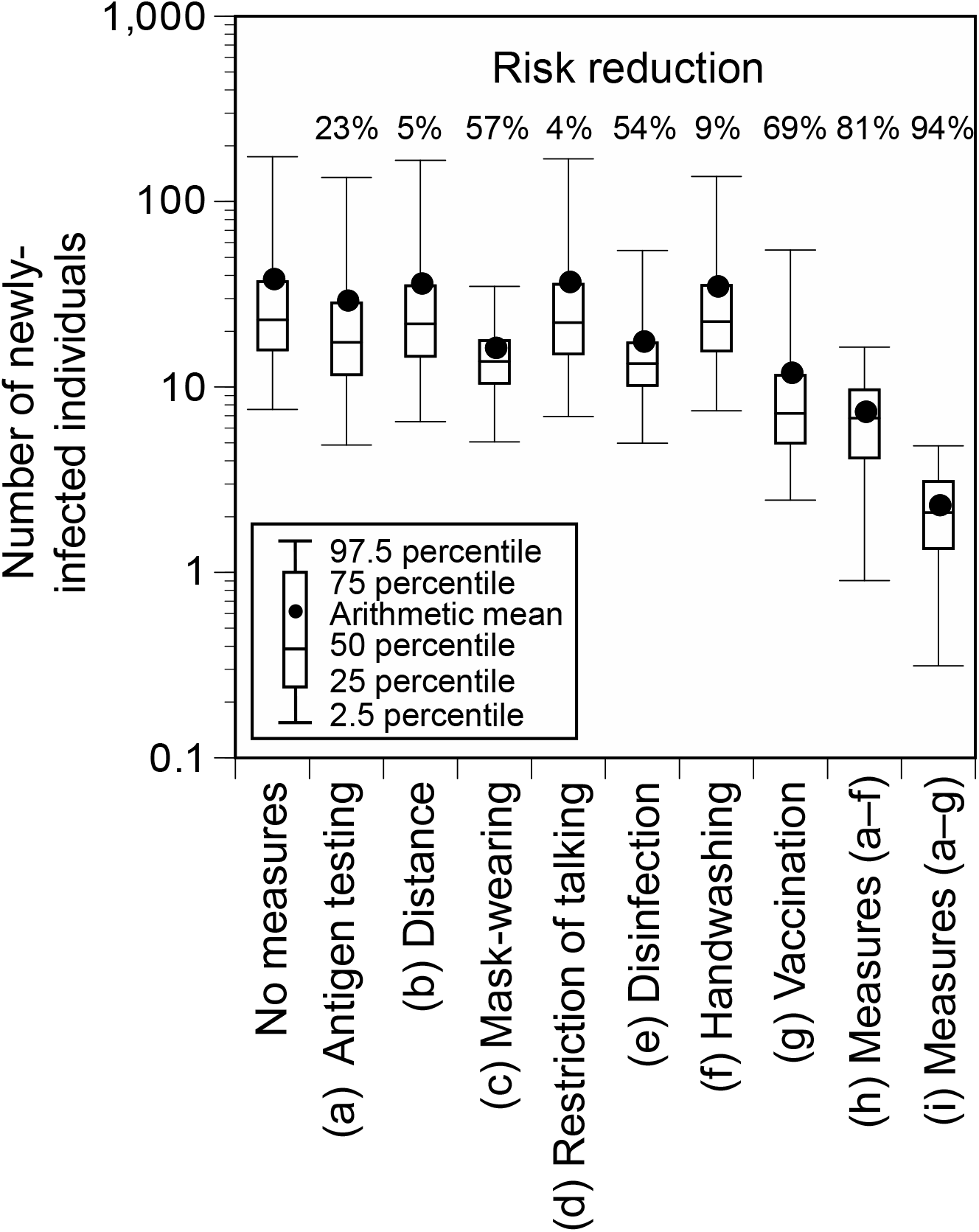
Number of newly-infected individuals and risk reduction when additional measures were applied to the base scenario. Viral concentration in the saliva: 100-fold increase relative to the wild-type strain.

When additional measures were implemented individually, the number of newly-infected individuals significantly decreased by vaccination (69%), mask-wearing (57%), and disinfection (54%), and the risk of infection was greatly reduced by implementing all the control measures (all measures except for vaccination: 81%; all measures including vaccination: 94%). When all measures, except for vaccination, were implemented and the number of participants or *P*_0_ was reduced, the number of newly-infected individuals was linearly related to the reduction ratio of the number of participants or *P*_0_ (Figure S2). If the event organizer considered keeping the number of newly-infected individuals below five as the arithmetic mean and below 10 as the 97.5 percentile, the number of participants or *P*_0_ had to be less than or equal to 50% of the base scenario.

## DISCUSSION

In this study, the number of infected individuals was estimated using an environmental exposure model for an outdoor music festival during which the cluster outbreak occurred. The risk reduction by the implementation of additional control measures was evaluated. The reported value was in the range of 95% UI of the total estimated number of infected individuals at any condition (10-, 100-, and 1,000-fold increase of the Delta-variant concentrations relative to the wild-type strain). It agreed well with the arithmetic mean of the values obtained for the condition with the 100-fold increase in the viral concentration. The results of the sensitivity analysis with varying mask-wearing proportions also showed that the reported value was within the range of the estimates. The reported number of infected individuals might have been underestimated because not all the participants were tested.

Based on the information from the free PCR testing that was conducted in the Aichi Prefecture and Nagoya City (eight positive cases among 658 people ^10^), the number of infected individuals was determined to be 90. This value was within the 95% UI of the number of infected individuals under conditions in which the viral concentration was 100 or 1,000 times higher. Considering that the viral loads of the Delta strain are approximately 1,000 times higher than those of the wild-type strain ^18^, these results support the validity of the infection risk assessment using the environmental exposure model.

Subsequently, we evaluated the extent to which the risk could be reduced by strengthening the infection control measures. Among the additional individual measures, vaccination, mask-wearing, and disinfection of surfaces were effective. The combination of all measures resulted in a higher risk reduction (all measures excluding vaccination: 81%; all measures including vaccination: 94%). Thus, the infection risk can be reduced by blocking all pathways of virus transmission including direct exposure, direct inhalation, contact, and air inhalation.

During mass gathering events, the extent to which any measures are implemented depends on the organizers’ decisions or society’s consensus on how many newly-infected individuals are acceptable. For example, in this study, the number of newly-infected individuals was estimated to be 7.2 (95% UI: 0.9–16.4) even if all measures, except for vaccination, were implemented. If the benchmark of acceptable newly-infected individuals was set to less than five and 10 as the arithmetic mean and 97.5 percentile, respectively, additional measures would be necessary such as allowing only vaccinated people to participate or limiting the number of participants to less than or equal to 50%. In addition, although the infection status fluctuates from time to time, there is a linear relationship between *P*_0_ and the number of newly-infected individuals, which makes it possible to determine whether additional measures are necessary for holding mass gathering events or whether to refrain from holding such events.

This study has several limitations. First, the risk of infection outside the event was not assessed in this study; however, confirmed infected individuals may have been infected during activities outside the event. In particular, those who accompany infectors might also act together, even outside the event. Second, we validated the model based on the total number of infected individuals but did not validate the detailed calculations within the model such as the exposure rates related to each infection pathway and the risk of infection for each type of exposed person. Case-control studies with behavioral records of event participants and environmental measurements of viral concentrations in the air and surface would fill these knowledge gaps.

Despite these limitations, a model for outdoor music festivals was successfully developed in this study and its validity was evaluated. The results of this study guide decision-making related to event organization such as the need to implement additional measures.

## Data Availability

All data produced in the present work are contained in the manuscript.

## ACKNOWLEDGEMENTS

We would like to thank Editage (www.editage.com) for English language editing and Dr. Kotoe Katayama and Dr. Masaaki Kitajima for their advice.

## DECLARATION OF INTERESTS

Support for the reported work:

This research received no external financial or non-financial support.

### Relevant support outside this work

T.Y. reports a relationship with Kao Corporation, Nippon Professional Baseball Organization, Yomiuri Giants, Tokyo Yakult Swallows, the Japan Professional Football League, and the Japan Professional Basketball League: funding grants.

### Intellectual property

There are no patents to disclose.

### Other activities

This study was conducted as part of a comprehensive research project, comprising members from two private companies, Kao Corporation and NVIDIA Corporation, Japan. No authors in this study belong to these companies. M.M., S.I., and T.Y. have attended the new coronavirus countermeasures liaison council jointly established by the Nippon Professional Baseball Organization and Japan Professional Football League as experts without any rewards. T.Y. is an advisor to the Japan National Stadium and Japan Professional Football League. Other authors declare no competing interests. The findings and conclusions of this article are solely the responsibility of the authors and do not represent the official views of any institution.

## AUTHORSHIP CONTRIBUTION

**Michio Murakami**: Conceptualization; Data curation; Methodology; Visualization; Project administration; Writing –original draft

**Tsukasa Fujita**: Formal analysis; Methodology; Writing –review & editing

**Pinqi Li**: Formal analysis; Methodology; Writing –review & editing

**Seiya Imoto**: Project administration; Supervision; Writing –review & editing

**Tetsuo Yasutaka**: Methodology; Project administration; Writing –review & editing

## Supporting Information

**Table S1.**
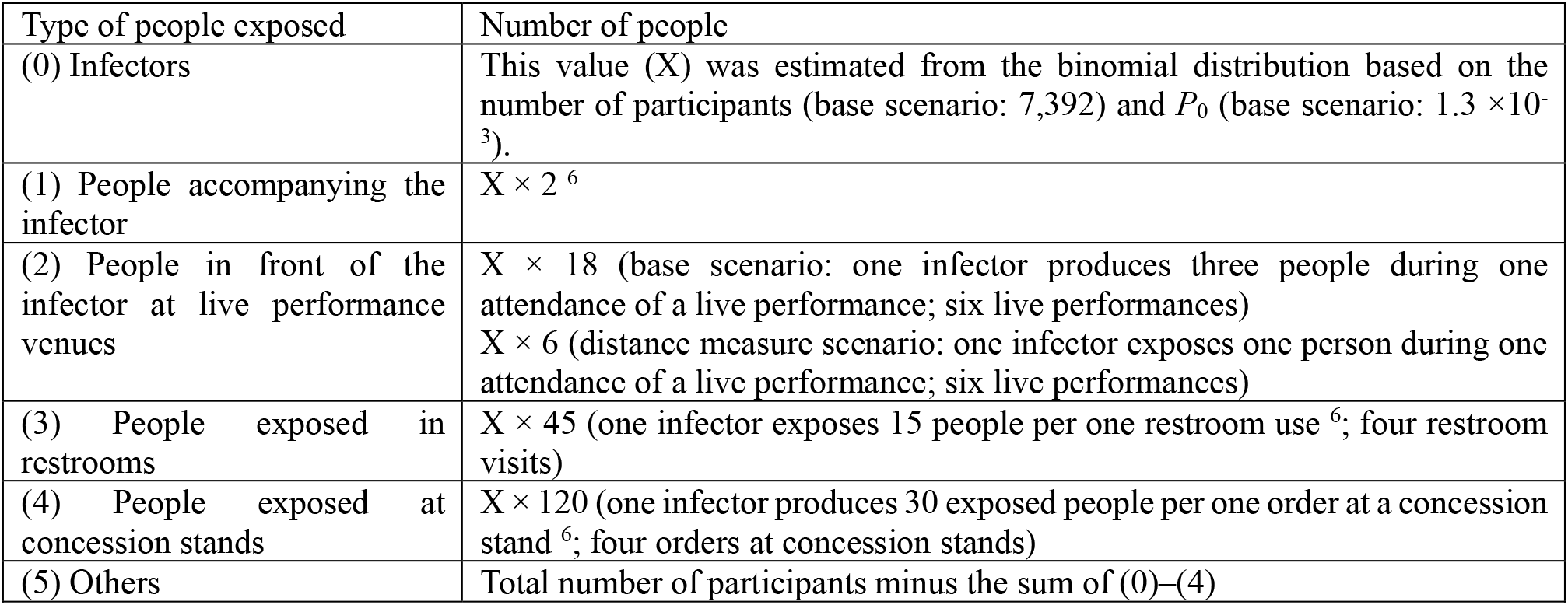
Type and number of people exposed. *P*_0_: crude probability of a participant being an infector.

**Table S2.** Pathways of infection by behavioral pattern.

| Behavioral pattern | Type of people exposed | Pathway | Note |
| --- | --- | --- | --- |
| (A) Attending live performances | People accompanying the infector | Direct droplet spray, direct inhalation of inspirable particles, and inhalation of respirable particles via air | The distance between the infector and the accompanying people or people in front of the infector was as follows: 0.5 m (base scenario), 1 m (distance measure scenario)<br>Frequency of talking of the infector: 0.2 per minute (base scenario), 0.03 per minute (talk measure scenario)<br>The probability that an infector faces each accompanying person and the people in front was 15% and 70%, respectively.<br>The probability that the accompanying person faces the infector was 50%. |
|  | People in front of the infector at live performance venues | Direct inhalation of inspirable particles and inhalation of respirable particles via air |  |
|  | People exposed in restrooms, people exposed at concession stands, and others | Inhalation of respirable particles via air |  |
| (B) Entering, exiting, and resting | People accompanying the infector | Direct droplet spray, direct inhalation of inspirable particles, and inhalation of respirable particles via air | The distance between the infector and the companions was as follows: 0.5 m (base scenario), 1.5 m (distance measure scenario)<br>Frequency of talking of the infector: 0.2 per minute<br>The probability that an infector faces each accompanying person was 50%.<br>The probability that the accompanying person faces the infector was 50%. |
|  | People in front of the infector at live performance venues, people exposed in restrooms, people exposed at concession stands, and others | Inhalation of respirable particles via air |  |
| (C) Using restrooms | People exposed in restrooms | Hand contact | The person touches the contaminated surface two minutes after the virus was deposited on the surface. The exposure from fingers-to-face contact was considered to be 6 h.<br>Frequency of talking of the infector: 0 per minute.<br>Handwashing measures inactivate the virus on fingers.<br>Wearing a mask reduces the frequency of touching the facial mucosal membranes. |
| (D) Ordering at concession stands | People exposed at concession stands | Hand contact | The person touches the contaminated surface 1 min after the virus was deposited on the surface. The exposure from fingers-to-face contact was considered to be 6 h.<br>Frequency of talking of the infector: 1 per minute. By considering the talk time to be 10 s, the amount of virus emitted by talking was assumed to be 1/6<sup>th</sup> of that per minute.<br>Disinfection measures inactivate the virus on surfaces.<br>Wearing a mask reduces the frequency of touching the facial mucosal membranes. |
| (E) Eating | People accompanying the infector | Direct droplet spray, direct inhalation of inspirable particles, and inhalation of respirable particles via air | The distance between the infector and the accompanying people was as follows: 0.5 m (base scenario), 1.5 m (distance measure scenario)<br>Frequency of talking of the infector: 0.2 per |
|  | People in front of the infector at live performance venues, people exposed in restrooms, people exposed at concession stands, and others | Inhalation of respirable particles via air | minute (base scenario), 0.03 per minute (talk measure scenario)<br>The probability that an infector faces each accompanier was 50%.<br>The probability that the accompanying person faces the infector was 50%.<br>People do not wear masks during meals. |

**Table S3.** Dose by type of person exposed.

| Types of people exposed | Dose |
| --- | --- |
| (1) People accompanying the infector | (A) Attending live performances: (direct droplet spray + direct inhalation of inspirable particles + inhalation of respirable particles via air) $\times 6$<br>(B) Entering, exiting, and resting: (direct droplet spray + direct inhalation of inspirable particles + inhalation of respirable particles via air) $\times 6$<br>(E) Eating: (direct droplet spray + direct inhalation of inspirable particles + inhalation of respirable particles via air) $\times 2$ |
| (2) People in front of the infector at live performance venues | (A) Attending live performances: (direct inhalation of inspirable particles) $\times 1$ + (inhalation of respirable particles via air) $\times 6$<br>(B) Entering, exiting, and resting: (inhalation of respirable particles via air) $\times 6$<br>(E) Eating: (inhalation of respirable particles via air) $\times 2$ |
| (3) People exposed in restrooms | (A) Attending live performances: (inhalation of respirable particles via air) $\times 6$<br>(B) Entering, exiting, and resting: (inhalation of respirable particles via air) $\times 6$<br>(C) Using restrooms: (hand contact) $\times 1$<br>(E) Eating: (inhalation of respirable particles via air) $\times 2$ |
| (4) People exposed at concession stands | (A) Attending live performances: (inhalation of respirable particles via air) $\times 6$<br>(B) Entering, exiting, and resting: (inhalation of respirable particles via air) $\times 6$<br>(D) Ordering at concession stands: (hand contact) $\times 1$<br>(E) Eating: (inhalation of respirable particles via air) $\times 2$ |
| (5) Others | (A) Attending live performances: (inhalation of respirable particles via air) $\times 6$<br>(B) Entering, exiting, and resting: (inhalation of respirable particles via air) $\times 6$<br>(E) Eating: (inhalation of respirable particles via air) $\times 2$ |

**Figure S1.**
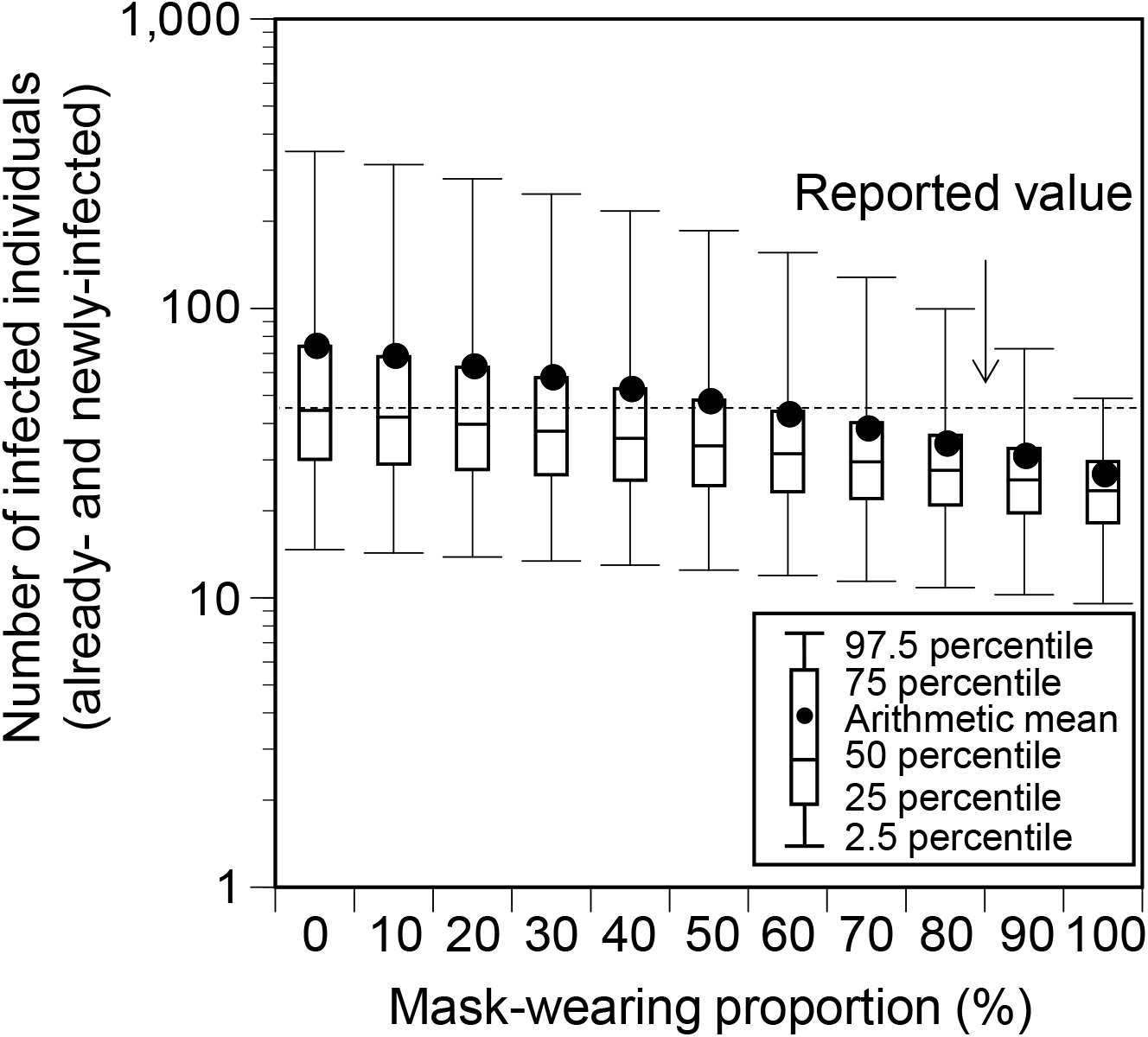
Comparison of the estimated and reported numbers of already- and newly-infected individuals under conditions with varying mask-wearing proportions. Viral concentration in the saliva: 100-fold increase relative to the wild-type strain. No additional measures (base scenario).

**Figure S2.**
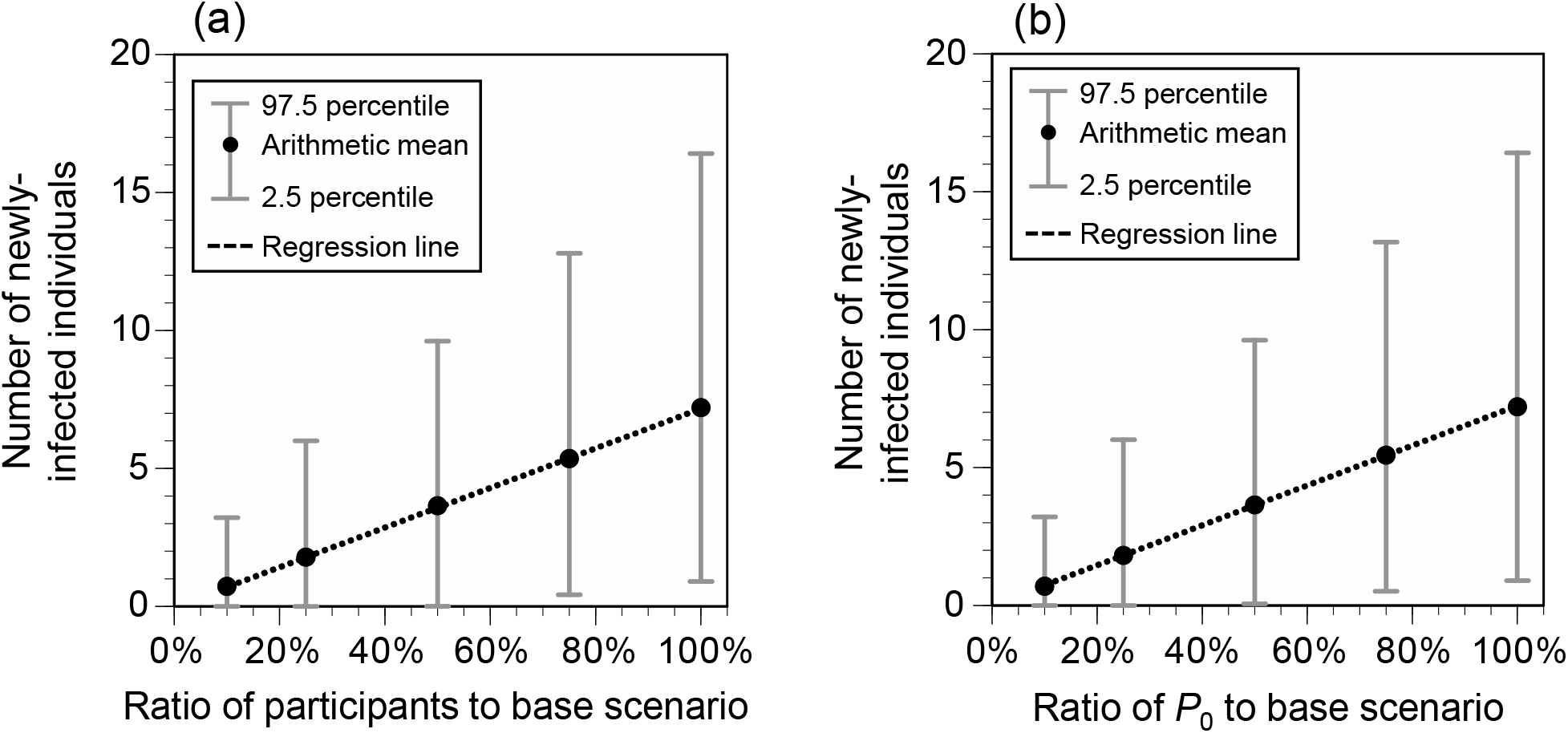
Number of newly-infected individuals for varying ratios of the number of participants (a) and *P*_0_ (b) to the base scenario. *P*_0_: crude probability of a participant being an infector. Viral concentration in the saliva: 100-fold increase relative to the wild-type strain. Additional measures (a–f) were implemented. When the number of participants was 10% (739), the sum of infectors, people accompanying the infector, people in front of the infector at live performance venues, people exposed in restrooms, and people exposed at concession stands exceeded the number of participants in seven of 10,000 simulations. The number of newly-infected individuals in these runs was calculated by summing the number of newly-infected individuals calculated for each group and dividing it by the total number of participants (739).

